# Accessible and reliable neurometric testing in humans using a smartphone platform

**DOI:** 10.1101/2023.02.01.23285291

**Authors:** H.J. Boele, C. Jung, S. Sherry, L.E.M. Roggeveen, S. Dijkhuizen, J. Öhman, E. Abraham, A. Uvarov, C.P. Boele, K. Gultig, A. Rasmussen, M.F. Vinueza-Veloz, J.F. Medina, S.K.E. Koekkoek, C.I. De Zeeuw, S.S.-H. Wang

**Affiliations:** Princeton Neuroscience Institute, Princeton, USA; Department of Neuroscience, Erasmus MC, Rotterdam, The Netherlands; Department of Clinical Sciences, Lund University, Sweden; BlinkLab Pty LTD, Sydney, Australia; Department of Neuroscience, University of Amsterdam, The Netherlands; Department of Community Medicine and Global Health, University of Oslo, Oslo, Norway; Department of Neuroscience, Baylor College of Medicine, USA; Netherlands Institute for Neuroscience, Royal Academy of Arts and Sciences, Amsterdam

**Keywords:** Eyeblink conditioning, Prepulse inhibition, Startle habituation, Smartphone-based, Neurobehavioral testing

## Abstract

Tests of human brain circuit function typically require fixed equipment in lab environments. We have developed a smartphone-based platform for neurometric testing. This platform, which uses AI models like computer vision, is optimized for at-home use and produces reproducible, robust results on a battery of tests, including eyeblink conditioning, prepulse inhibition of acoustic startle response, and startle habituation. This approach provides a scalable, universal resource for quantitative assays of central nervous system function.

## INTRODUCTION

Neurobehavioral assays of brain function can reveal fundamental mechanisms underlying neuropsychiatric conditions (1,2), but typically require centrally located equipment in a laboratory test facility. Consequently, these tests are often unpleasant for participants as they require instruments attached to their face and cannot be used at scale in daily clinical practice.

We have developed a smartphone-based software platform, termed BlinkLab, to perform neurobehavioral testing free from facial instruments or other fixed-location equipment. This AI platform is designed to be used at home or in similar environments, independently or with the assistance of a caregiver, while following instructions from the mobile-device application. The tests include, but are not limited to, eyeblink conditioning, a form of sensory-motor associative learning, prepulse inhibition of the acoustic startle response, which measures the ability to filter out irrelevant information through sensorimotor gating, and startle habituation, which measures the ability for the intrinsic damping of repetitive stimuli.

The BlinkLab application combines a smartphone’s ability to deliver stimuli and acquire data using computer vision with a secure cloud-based portal for data storage and analysis (**Fig. 1**). In our experiments, each audio and/or visual stimulus is presented with millisecond-precise control over parameters such as timing, amplitude and frequency. In order to maintain participant attention, an entertaining movie of choice is shown with normalized audio levels. Participants’ responses are measured by the smartphone’s camera and microphone, and are processed in real time using state-of-the art computer vision techniques, fully anonymized, and transferred securely (TLS 1.3) to the analysis portal.

**Figure 1.**
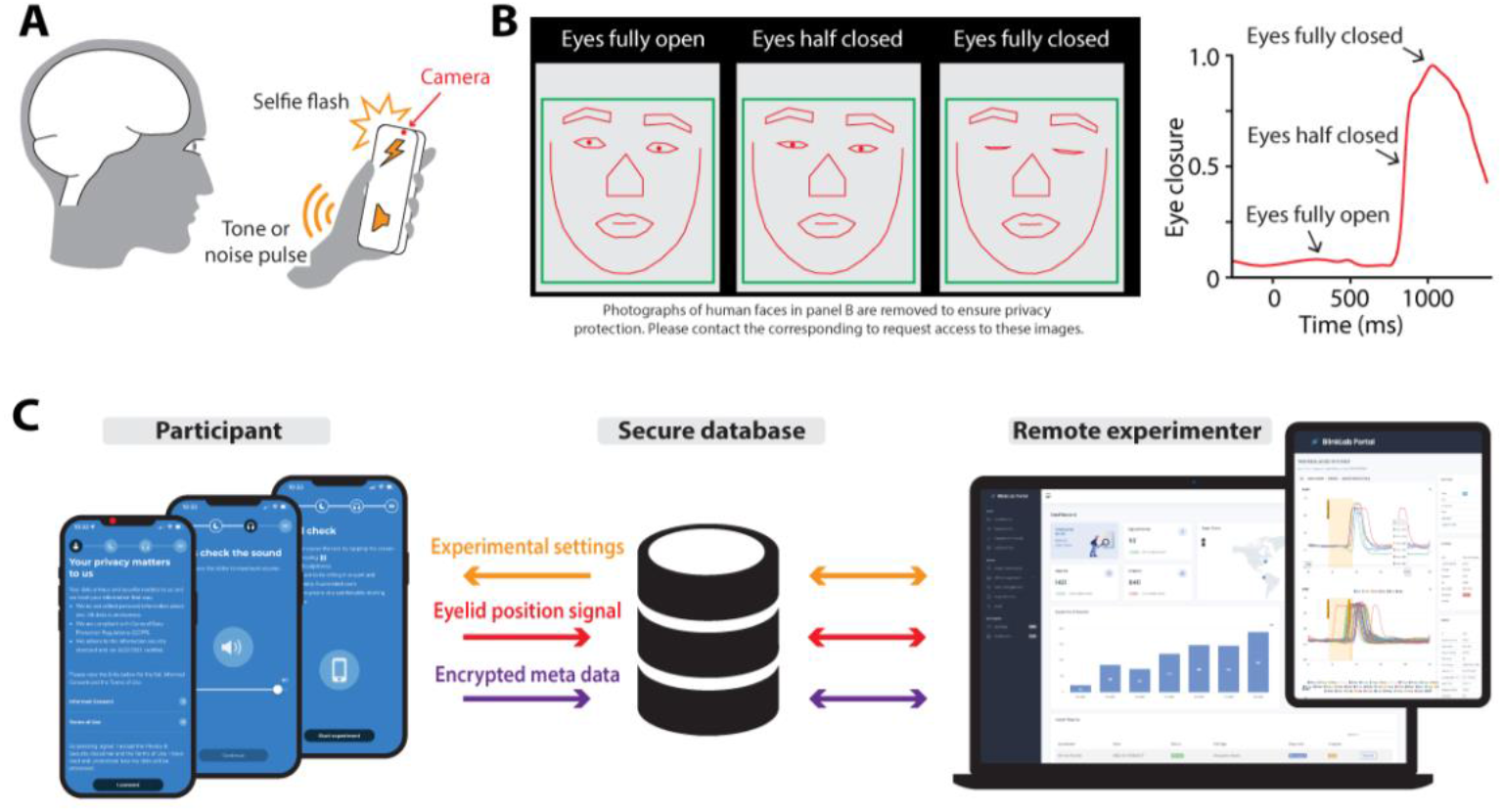
Architecture of smartphone-mediated neurobehavioral testing. **(A)** Auditory, visual, and tactile stimuli are delivered via the smartphone. The camera measures the participant’s responses at 60 Hz. **(B)** Facial landmark detection algorithms are capable of detecting eyelid movements in real-time on the smartphone. Images of the face are removed to protect privacy. Contact corresponding author to request access to these images. Images are used and can be shared with permission of the participant. **(C)** The architecture of smartphone-mediated neurobehavioral testing includes a smartphone application (left), a secure database (middle), and a cloud-based analysis portal (right) that allows the remote experimenter to control experimental parameters and analyze collected data.

## MATERIALS AND METHODS

### Subjects/Participants

Participants were recruited from Princeton University (United States) and Erasmus MC (The Netherlands). For Princeton University, participants were invited using the SONA system. For Erasmus MC, participants were recruited using flyers and invites. Participants were informed of the institutional guidelines, to which they gave their written consents and permissions. There were no restrictions placed on sex at birth, gender they identify with, nor race. Excluded were participants younger than 12 and those formally diagnosed with a neurodevelopmental, neuropsychiatric, or neurodegenerative condition. All procedures were approved by both the Institutional Review Board for Human Subjects of Princeton University (IRB#13943) and the Medical Ethics Review Committee of Erasmus MC (# MEC 2022 0116).

### Experimental setup

The smartphone application was developed in Swift, a compiled programming language developed by Apple. The cloud-based analysis portal was developed in Symfony, a high-performance PHP framework for web development which uses PostgreSQL 14 as the database. The smartphone application was distributed using TestFlight and the AppStore. Eyelid movements were recorded with the smartphone’s front-facing camera at 60 frames per second. All stimuli were controlled by the BlinkLab app. The models of iPhones used were iPhone X, iPhone 11, iPhone 13 Pro, and iPhone 13 Pro Max. The headphones used were Pioneer over-ear wired headphones. The total length of a session was approximately 15 minutes for a single eyeblink conditioning session, 12 minutes for a prepulse inhibition session, and 5 minutes for a startle habituation session (actual length depended on the total durations of the random intertrial intervals).

Prior to the start of the test, we assured that the participant was in a quiet environment with ambient lighting and in a comfortable position. The app continuously monitored the surrounding environment, including the light intensity using the camera, the background noise using the microphone, and the smartphone position and movements using the accelerometer. If one of the aforementioned values was out of bounds, the video paused and the app instructed the user on how to change the environment or try taking the test at another time. During the experiment, the user watched an entertaining (audio normalized) movie while the stimuli for eyeblink conditioning, prepulse inhibition, or startle habituation were delivered. For each trial, facial landmark detection algorithms were used to track and record the position of the participant’s facial landmarks over time to determine amplitude and timing of the eyelid closure. Users could see a small progress bar at the bottom of the screen that showed them how far along they were in the experiment. Results were securely transferred and stored in a cloud-based analysis environment where data became immediately accessible for researchers through the BlinkLab analysis portal. Both raw and processed data is available in the most widely used data standards, as well as through BlinkLab’s cloud based analysis and visualization tools.

## RESULTS

Eyeblink conditioning using the smartphone approach induced conditioned responses comparable with traditional stimuli such as an airpuff^18^. The unconditioned stimulus (US) was a 50 ms white noise pulse paired with a brief screen flash, which reliably elicited a reflexive eyelid closure and activated cerebellum-dependent learning mechanisms (3,4). The conditioned stimulus (CS) was a white 1 cm circular dot presented for 450 ms as an overlay over the movie at the screen’s center **(See online materials)**.

Repeated pairings of the CS and US in a delay paradigm on the smartphone, with an interstimulus interval (ISI) of 400 ms, resulted in a robust acquisition of eyelid conditioned responses (CR) at the end of six sessions (acquisition phase) of fifty paired CS-US trials each. The CR amplitude increased from -0.02 (± 0.05 95% CI) in baseline session 0 to 0.35 (± 0.09 95% CI) in session 6 (F (6,4044) = 74.82, p < .001, linear mixed model (LME)) **(Fig. 2A, Table S2)**. Similarly, the CR percentage increased from 5.3 (± 6.6 95% CI) in baseline session 0 to 58.5 (± 10.6 95% CI) in session 6 (F (6,4044) = 65.13, p < .001, LME)) (**Fig. 2B, Table S2**). CR timing significantly improved over the course of training, with session 6 yielding CRs with a latency to peak around the onset of the expected US at 470.46 (± 26.53 95% CI) ms after CS onset (F (6,270) = 8.92, p < .001, ANOVA on LME). To confirm the putative cerebellar nature of these conditioned responses, we performed two additional tests after the acquisition phase. First, we tested the effect of a probe CS that was relatively short (i.e., only 100 ms) compared to the one that was used during training (session 7). This probe CS was only presented in CS-only trials and never reinforced with a US. In line with previous reports (5,6), we found that this short probe CS was able to elicit normal CRs in terms of CR percentage (F(1,1169) = 0.57, p = 0.450) and CR amplitude (F (1,1169) = 0.68, p = 0.41, LME) (**Fig 2C, D, Table S2**). Second, we tested the effect of an extension of the interstimulus interval from 400 ms to 700 ms during two additional training sessions (sessions 8 and 9). We found that the timing of eyeblink CRs adapted to the new longer ISI with a new latency to CR peak of 765.6 (± 75.85 95% CI) ms (**Fig 2C, E, Table S2**) (F (2,113) = 42.21, p < .001, ANOVA on LME), consistent with previous studies on eyeblink conditioning (7). As a result of our smartphone approach, we were able to induce robust learning in the eyeblink conditioning paradigm and produce data that had less variability in learning curves previously reported, including in our own studies on human eyeblink conditioning (compare for our smartphone approach with for instance (8–10)). The overall learning pattern now closely resembles those described previously in mouse and rabbit eyeblink conditioning literature (11–13), with a smooth, gradual increase of the CR amplitude, percentage, and timing that is adaptive to the interstimulus interval.

**Figure 2.**
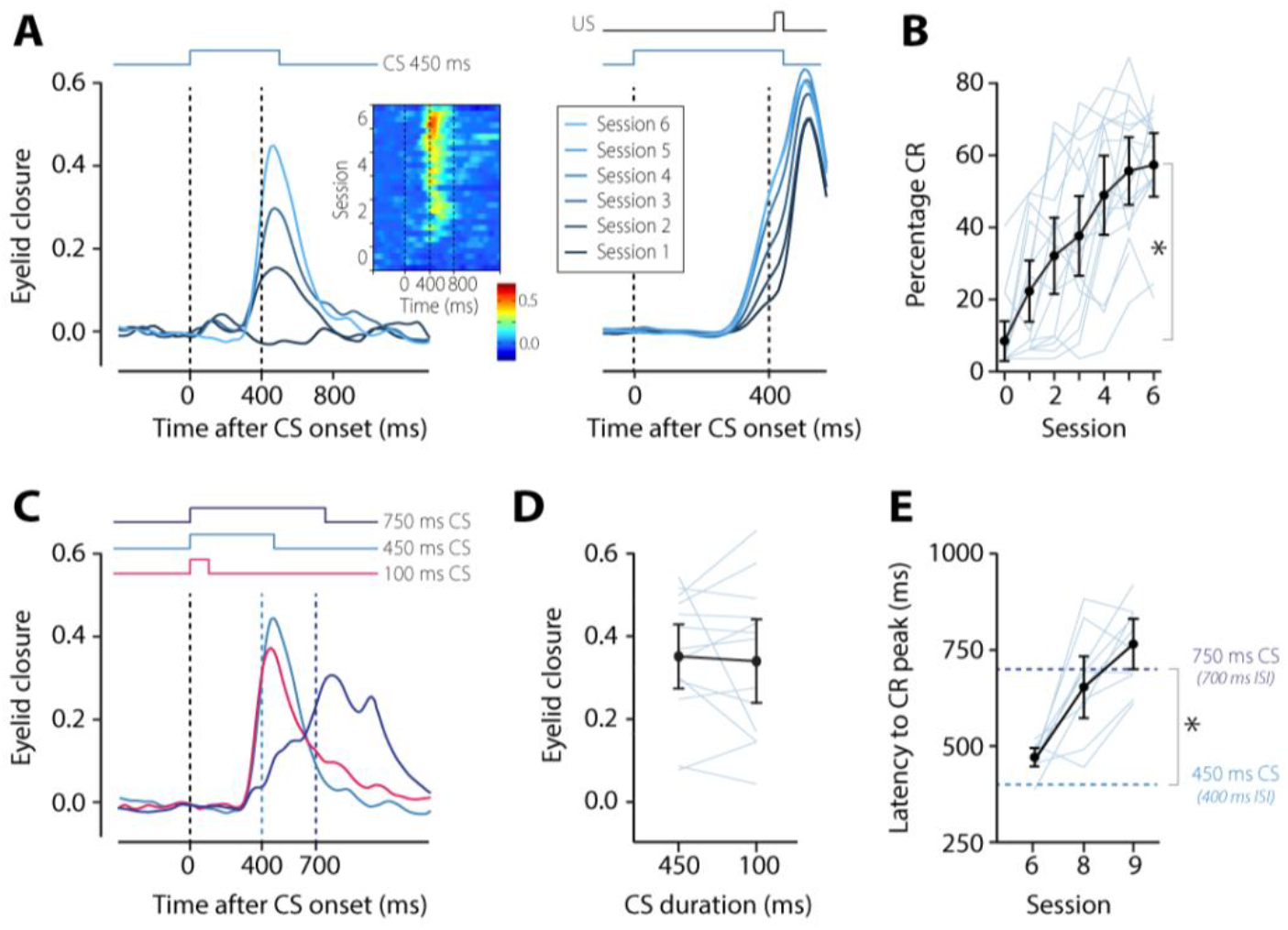
Smartphone-mediated delay eyeblink conditioning. **(A)** Session averaged eyeblink traces of CS-only trials (left) and paired CS-US trials (right). Note the gradual acquisition of eyeblink conditioned responses. The first vertical dashed line indicates the onset of the CS, the second one indicates the onset of the US. The heatmap shows the average per trial for CS-only trials during the 6 acquisition sessions; heat represents the amplitude of eyelid response. **(B)** Percentage of CRs per session. Light blue lines are individual learning curves, the thick black line represents the group average. **(C)** Session averaged eyelid CRs in response to a CS with a duration of 450 ms (light blue) at the end of acquisition (session 6), a short probe CS with a duration of 100 ms (brown), and a long CS with a duration of 750 ms (dark blue). The short CS was never reinforced with the US. The long CS was reinforced with a US. **(D)** The short CS of 100 ms elicited eyeblink CRs that were indistinguishable from those evoked by the original 450 ms CS. **(E)** As a result of the ISI switch, the latency to CR peak shifted from the onset of the old US at 400 ms to the onset of the new US at 700 ms. Abbreviations: 100 ms CS, conditional stimulus with a duration of 100 ms; 450 ms CS, conditional stimulus with a duration of 450 ms (ISI 400 ms); 750 ms CS, conditional stimulus with a duration of 750 ms (ISI 700 ms); CR, conditioned response. All error bars represent 95% confidence intervals.

Next, we studied prepulse inhibition of the startle response, using a 50 ms white noise audio burst at 105 dB as the pulse and a similar burst ranging between 75 dB and 95 dB as the prepulse. The ISI was set at 120 ms **(See online materials)**. Eyelid startle responses in the pulse-only trials had an average amplitude of 0.35 (± 0.08 95% CI), while responses in trials where the pulse was preceded by the weaker prepulse ranged between 0.07 (± 0.03 95% CI) and 0.14 (± 0.06 95% CI) (**Fig. 2F, G, Table S3)**. We found a significant main effect of trial type (F (4,1434) = 99.44, p < .001, ANOVA on LME) and pairwise post-hoc testing revealed that the largest effects were present between the pulse and the prepulse + pulse trials (**Fig. 3, Table S3**). With prepulse inhibition values of about 70% (100-(0.10/0.35) * 100), our data shows a strong inhibition of the acoustic startle responses (compare our results for instance with: (14–16)).

**Figure 3.**
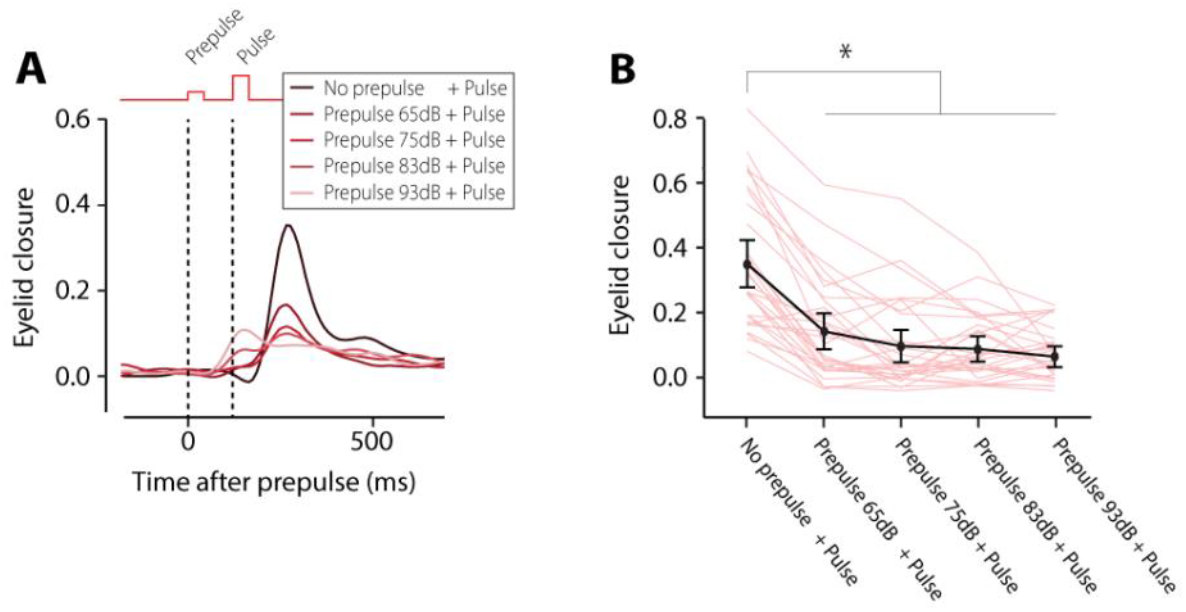
Smartphone-mediated prepulse inhibition of the acoustic startle response. **(A)** Averaged eyelid traces for the different trial types. The presentation of a soft sound (prepulse, first vertical dashed line) 120 ms before the loud sound (pulse, second vertical dashed line) resulted in a significant inhibition of the acoustic eyelid startle response. Note that higher prepulse intensities (25% and 50%) start to evoke a small startle response to the prepulse. **(B)** Amplitude of eyelid closure as a function of trial type. All error bars represent 95% confidence intervals.

Finally, we studied startle habituation using a 0.75 Hz pulse train of five white noise audio bursts at an intensity of 105 dB **(See online materials)**. We found a significant reduction of the eyelid startle amplitude over the course of the stimulus pattern, starting with 0.30 (± 0.15) at the first pulse and 0.13 (± 0.14) at the fifth pulse (**Fig. 4, Table S4**, F(4,612) = 18.14, p <.001, LME). Post-hoc testing revealed significant effects between pulse 1 and any of the other pulses (**Table S4**).

**Figure 4.**
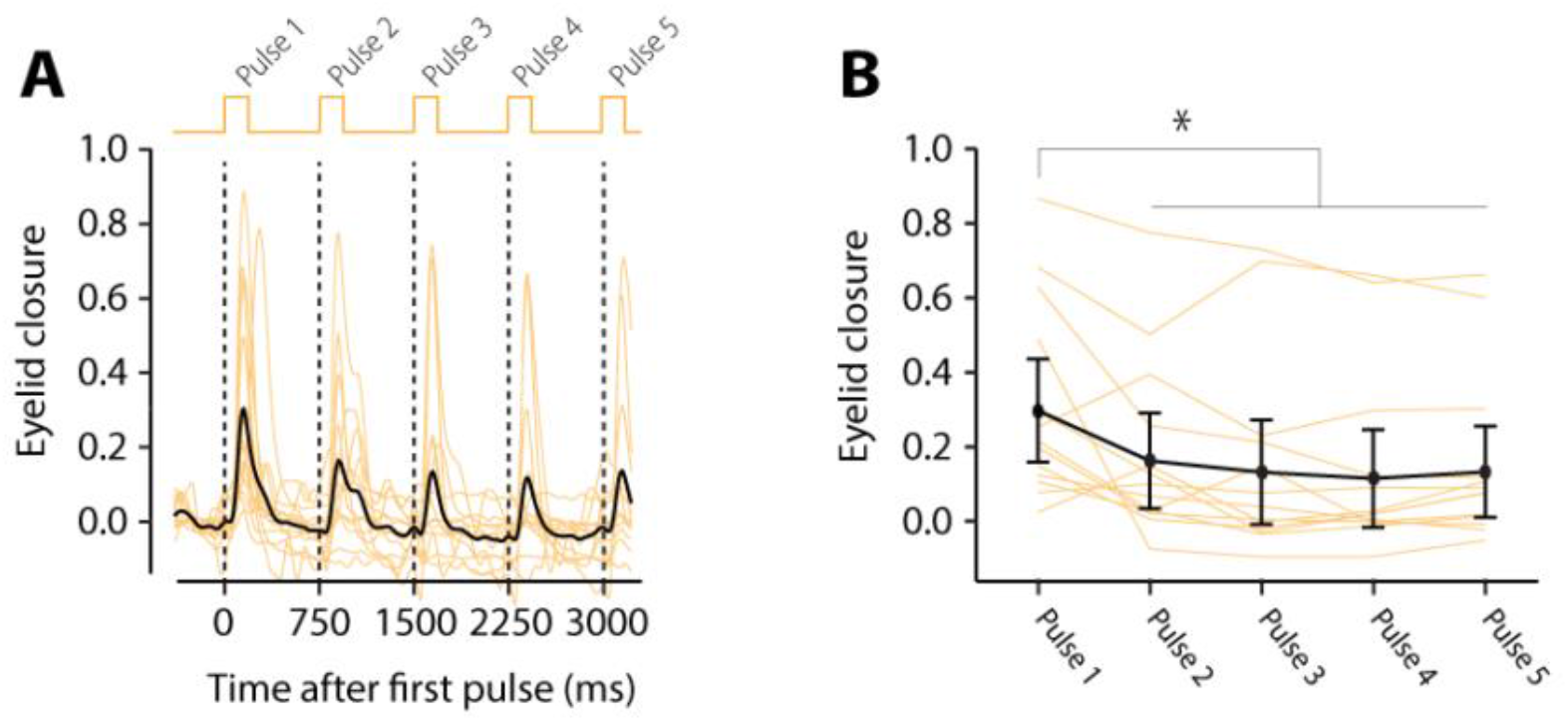
Smartphone-mediated startle habituation. **(A)** Individual (yellow) and group averaged (black) eyelid traces during the presentation of 5 consecutive acoustic white noise pulses, showing a gradual decline in eyelid startle responses. **(B)** Amplitude of eyelid closure as a function of trial type.

## DISCUSSION

Together, our data shows that we can now perform well-established neurobehavioral testing using accessible smartphone technology. In contrast to conducting these tests in a sterile laboratory environment, we found that people performed better and had less variability in their performance by doing them on a smartphone in a comfortable home-like environment. Since these tests are reflex-based and do not require verbal or social interaction, they allow large-scale cross-cultural human studies and a foundation on cross-species translational research. It has been shown that performance in eyeblink conditioning, prepulse inhibition, and startle habituation is strongly correlated with neuropsychiatric conditions, including autism(1,2,8), schizophrenia(17,18), dementia(8,19,20), Parkinson’s(21) and Huntington’s disease(21–23). As such, these tests have repeatedly been suggested as a potential biomarker to diagnose and monitor (pharmaceutical) intervention of neurodevelopmental and neurodegenerative conditions. Since the new smartphone approach does not require unpleasant in-lab testing, it opens up the possibility of using these quantitative tests in clinical practice.

## Data Availability

DATA AVAILABILITY Complete eyeblink conditioning dataset, prepulse inhibition dataset, and startle habituation dataset are publicly available at: https://github.com/506574657220426F656C650D/POC-datasets. Data is anonymized, the unique identifier for the different subjects is the subject_id. CODE AVAILABILITY Complete R syntax for data analysis is available at: https://github.com/506574657220426F656C650D/POC-datasets.

https://github.com/506574657220426F656C650D/POC-datasets

## ACKNOWLEDGEMENTS

We thank all the participants in our studies.

## CONFLICT OF INTEREST

HB, SK, CB, SW, AU, and CZ engage with BlinkLab Pty Ltd, the exclusive licensee of the technology, as co-founders and equity holders. The remaining authors declare no competing interests.

## AUTHOR CONTRIBUTIONS

Conceptualization: HB, SS, AU, CB, SK,

Methodology: HB, CJ, SS, AU, CB, SK, KG

Investigation: HB, CJ, SS, LR, SD, JO, EA, SK, KG

Analysis: HB, CB, SK,

Visualization: HB, CB, SK,

Funding acquisition: HB, AU,

Project administration: HB, CJ, SS, LR,

Supervision: HB, AU, SK, CZ, SW

Writing – original draft: HB, CJ, SS, SK,

Writing – review & editing: HB, JM, CZ, SW, AR

## FUNDING

This work was financially supported by the Princeton Accelerator Award 2021 (HJB, SSH)

## DATA AVAILABILITY

Complete eyeblink conditioning dataset, prepulse inhibition dataset, and startle habituation dataset are publicly available at: https://github.com/506574657220426F656C650D/POC-datasets. Data is anonymized, the unique identifier for the different subjects is the subject_id.

## CODE AVAILABILITY

Complete R syntax for data analysis is available at: https://github.com/506574657220426F656C650D/POC-datasets.

## Supplementary materials for

### ONLINE METHOD SECTION

#### Eyeblink conditioning training paradigm

Participants (n=14, details in **Table S2**) completed 6-9 eyeblink conditioning sessions within a 14-day span, with no more than 2 sessions per day. The conditioned stimulus (CS) consisted of a white circular dot, 1 cm in diameter, in the middle of the screen which lasted for 450 ms. The unconditioned stimulus (US) consisted of a simultaneous 105 dB, 50 ms white noise pulse and a 50 ms full screen retina flash. The CS and US were presented in a delay paradigm, which means that the CS and US have a delay in onset, but temporally overlap and co-terminate at the end. For session 1-6, the interval between CS and US onset was set at 400 ms **(Supplementary figure 2)**. The interval between the trials was set randomly between 7 and 20 seconds. During sessions 1-6, participants received a total of 50 trials distributed over 5 blocks. Each block consisted of 1 CS-only, 1 US-only, and 8 paired CS-US trials, semi-randomly distributed throughout the block. In session 7, participants received a short CS (100 ms) in CS-only trials to demonstrate cerebellum-dependent response timing. In sessions 8 and 9, the duration of the interstimulus interval (ISI) was suddenly extended from 400 ms to 700 ms. The longer ISI of 700 ms was used to assess the level of response timing adaptability. A training session lasted for about 15-20 minutes.

#### Prepulse inhibition training paradigm

Participants (n = 30, details in **Table S2**) completed a single prepulse inhibition session. One session contained 55 trials. The two stimuli were defined as: 1) a pulse consisting of a 105 dB, 50 ms white noise audio burst and 2) a prepulse, consisting of a 50 ms white noise audio burst of varying amplitude that was always softer than the pulse. We used prepulse at four intensities: 65 dB, 75 dB, 83 dB and 93 dB. First, a total of 5 habituation trials containing white noise bursts of various soft intensities were presented. This allowed for the participant to relax and settle into the movie. After these 5 trials, 10 blocks of 5 trials were presented. Each block consisted of a pulse only trial and 4 prepulse-pulse trials with prepulse amplitudes of respectively 5, 10, 25, and 50% of the startle amplitude. A prepulse always preceded the pulse by 120 ms **(Supplementary Figure 3)**. Intertrial interval (ITI) was set at random between 10 and 25 seconds. A training session lasted for about 15-20 minutes.

#### Startle habituation training paradigm

Participants (n = 14, details in **Table S2**) completed a single startle habituation session. One session contained 10 trials. For startle habituation, we used a 0.75 Hz pulse train of five white noise audio bursts at an intensity of 105 dB **(Supplementary Figure 4)**. The ITI was set at random between 20 and 40 seconds. A training session lasted for about 15-20 minutes.

#### Data analysis

Individual eyeblink traces were analyzed with custom computer software (R Studio; Boston, MA, v1.3.1093). Eyelid position signals were calculated in real-time on the smartphone based on the xyz coordinate of the six landmarks around the eye. For this, we calculated the difference between the y position values of the sum of the two upper eyelid landmarks and the sum of the two lower eyelid landmarks with the x position values of the two eye corner landmarks (**Supplementary Figure 1)**. For each type of trial, a single snippet was taken from the video of the eyelid position signal hereafter called an ‘eyeblink trace’. Eyeblink traces were filtered in forward and reverse directions with a low-pass Butterworth filter using a cut-off frequency at 50 Hz. Trials were min-max normalized by aligning the 500 ms pre-stimulus baselines and normalizing the signal so that the amplitude of a full blink was 1 normalized eye closure.

This normalization was achieved by using spontaneous blinks as a reference. For each session, the maximum value was calculated and the individual traces were normalized by dividing each trace by this value. Therefore, in the normalized traces, a normalized eyelid closure of 1 corresponded with the eye being fully closed and a normalized eyelid closure of 0 corresponded with the eye being fully open.

#### Eyeblink conditioning

To quantify eyeblink conditioning, we used four outcome measures: (1) The CR amplitude, defined as the amplitude of the eyelid response in the 60 ms - US offset window; (2) the CR percentage, defined as the percentage of trials within a session that contained a CR, whereby a CR was defined as an eyelid movement larger than 0.15 and with a latency to CR peak between 60 ms after CS onset and US offset; (3) the latency to CR onset in ms after CS onset, and (4) the latency to CR peak in ms after CS onset. The calculation of the CR percentage, CR amplitude, and latency to CR onset included both paired CS-US trials and CS-only trials. For latency to CR peak, we only included CS-only trials since they show the full kinetic profile of the eyeblink CR and provide a better estimate of the adaptive timing of eyeblink CRs.

#### Prepulse inhibition

The response detection window for eyelid responses to the pulse was set at 60-330 ms after pulse onset. To analyze amplitude reduction, we used the amplitude of the normalized eyelid closure at the mean peak time of significant startle responses calculated over *all* trials. In addition to responsiveness to the startle stimuli, we analyzed the effect of the prepulse itself on normalized eyelid closure in a similar fashion but used a response window of 60-180 ms after the prepulse.

#### Startle habituation

The response detection windows for startle responses were set at 60-330 ms after a startle stimulus. Response detection was done in a similar fashion as described above for eyeblink conditioning. To analyze amplitude reduction, we used the amplitude of the normalized eyelid closure at the mean peak time of significant startle responses calculated over *all* trials.

#### Statistical analysis

Statistical analysis and data visualizations were done in R Studio (V2022.02.03) using the following packages: dplyr, emmeans, ggplot2, lmerTest, nlme, tidyr, and tidyverse. We used multilevel linear mixed-effects (LME) models in R Studio because they are more robust to violations of normality assumptions, which is often the case in biological data samples. LME models can better accommodate the nested structure of our data (i.e., trial nested within session, session nested within subject, subject nested within group) and prevent data loss by using summary measures. As an added benefit, LME models are better at handling missing data points than repeated measures analysis of variance (ANOVA) models and do not require homoscedasticity as an inherent assumption. In our LME, we used session as a fixed effect, and subject as a random effect. Goodness-of-fit model comparison was determined by evaluating log likelihood ratio, BIC, and AIC values. The distribution of residuals was inspected visually by plotting the quantiles of standard normal versus standardized residuals (*i*.*e*. Q-Q plots). Data were considered as statistically significant if the p-value was less than 0.05.

**Supplementary Figure 1.**
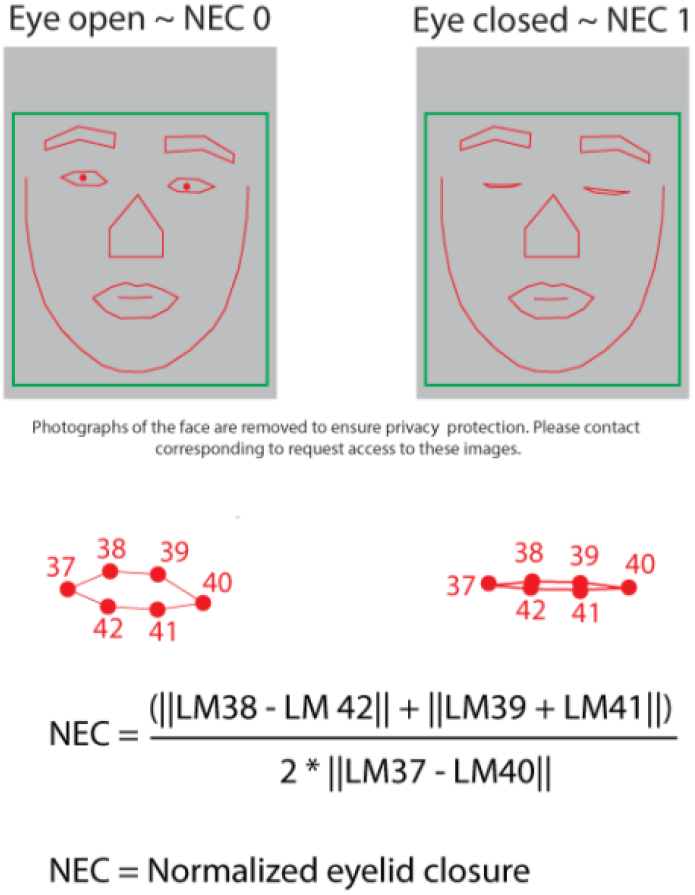
Computer vision algorithms were used to capture facial landmarks detection, including those of the eyelids. Normalized eyelid closure (NEC) was calculated for each eye, using six landmarks for each eye. Images of the face are removed to protect privacy. Contact corresponding author to request access to these images. Images were used and can be shared with permission of the participant.

**Supplementary Figure 2.**
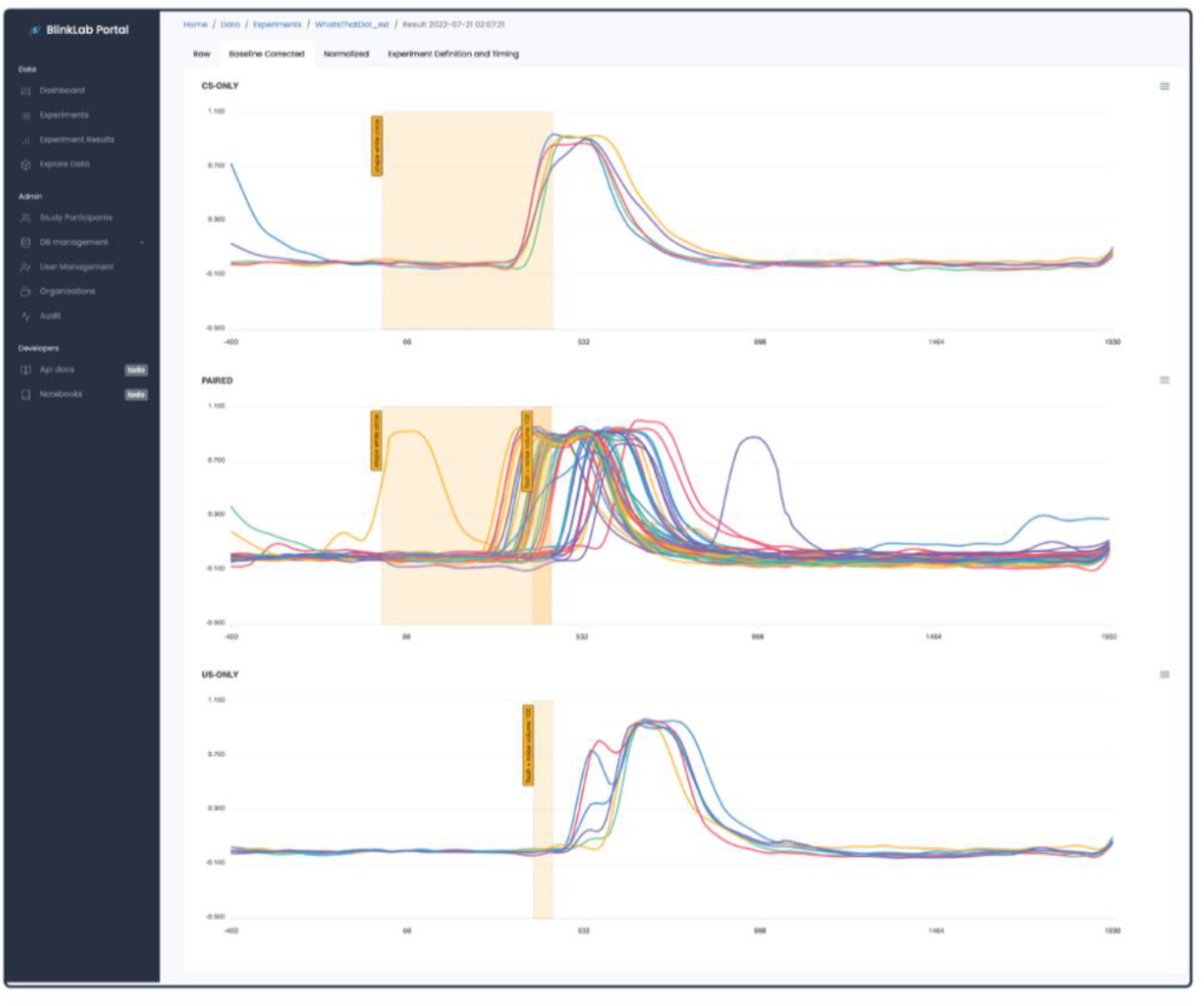
Raw eyelid traces of one participant for one eyelid conditioning session. This figure is a screen capture of the cloud-based analysis portal and shows how we visualize the data that is collected with the smartphone. Depicted is session 6, the last session of the acquisition phase. Each colored line represents one trial. Top panel, CS only trials; Middle panel, paired CS-US trials; Bottom panel, US only trials. Note the robust conditioned eyelid responses.

**Supplementary Figure 3.**
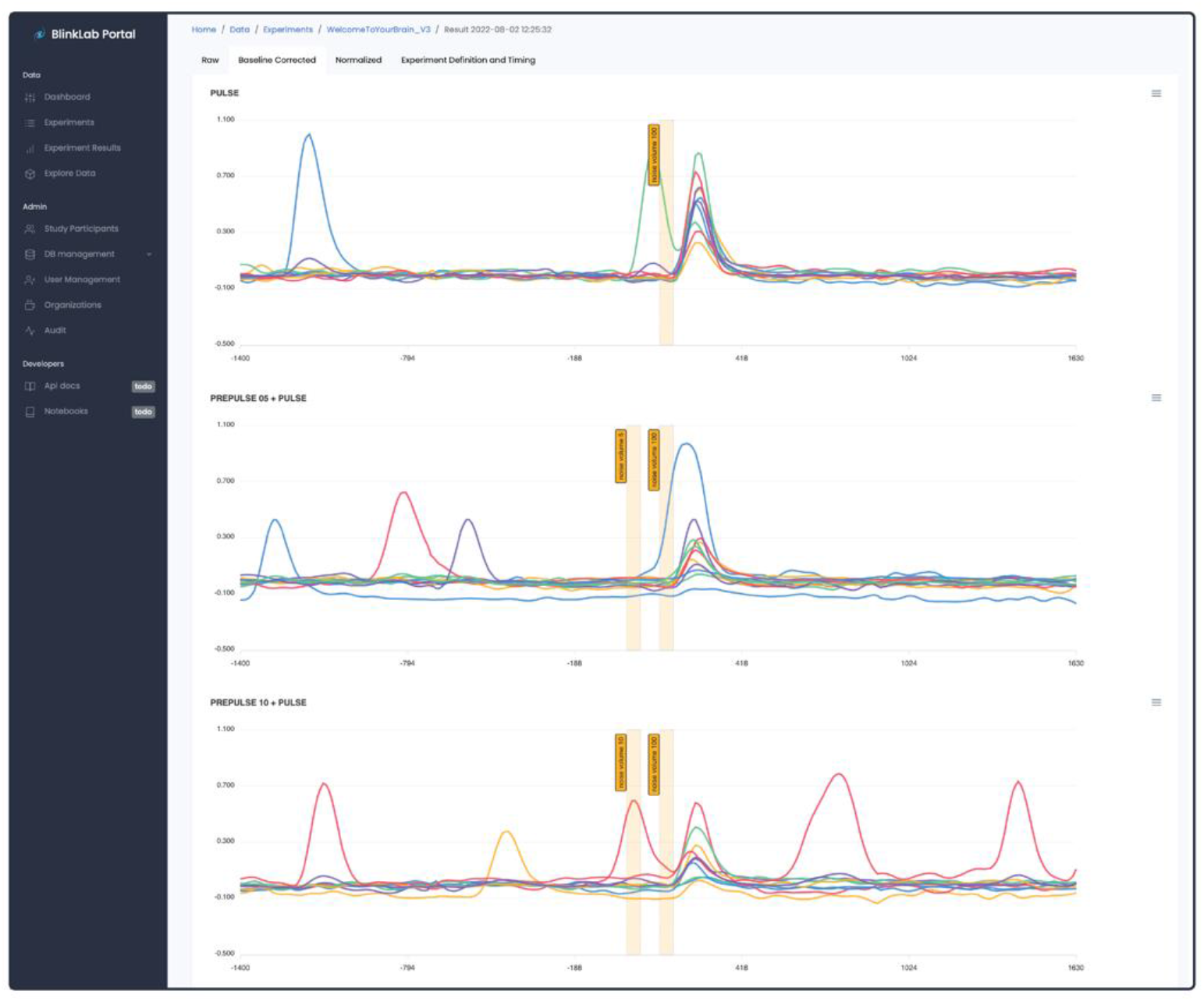
Raw eyelid traces of one participant for one prepulse inhibition of the acoustic startle experiment. This figure is a screen capture of the cloud-based analysis portal and shows how we visualize the data that is collected with the smartphone. Each colored line represents one trial. Top panel, pulse only trials (105 dB); Middle panel, prepulse + pulse trials (65 dB + 105 dB); Bottom panel, prepulse + pulse trials (75 dB + 105 dB). Note the inhibition of the eyelid startle response in the prepulse + pulse trials.

**Supplementary Figure 4.**
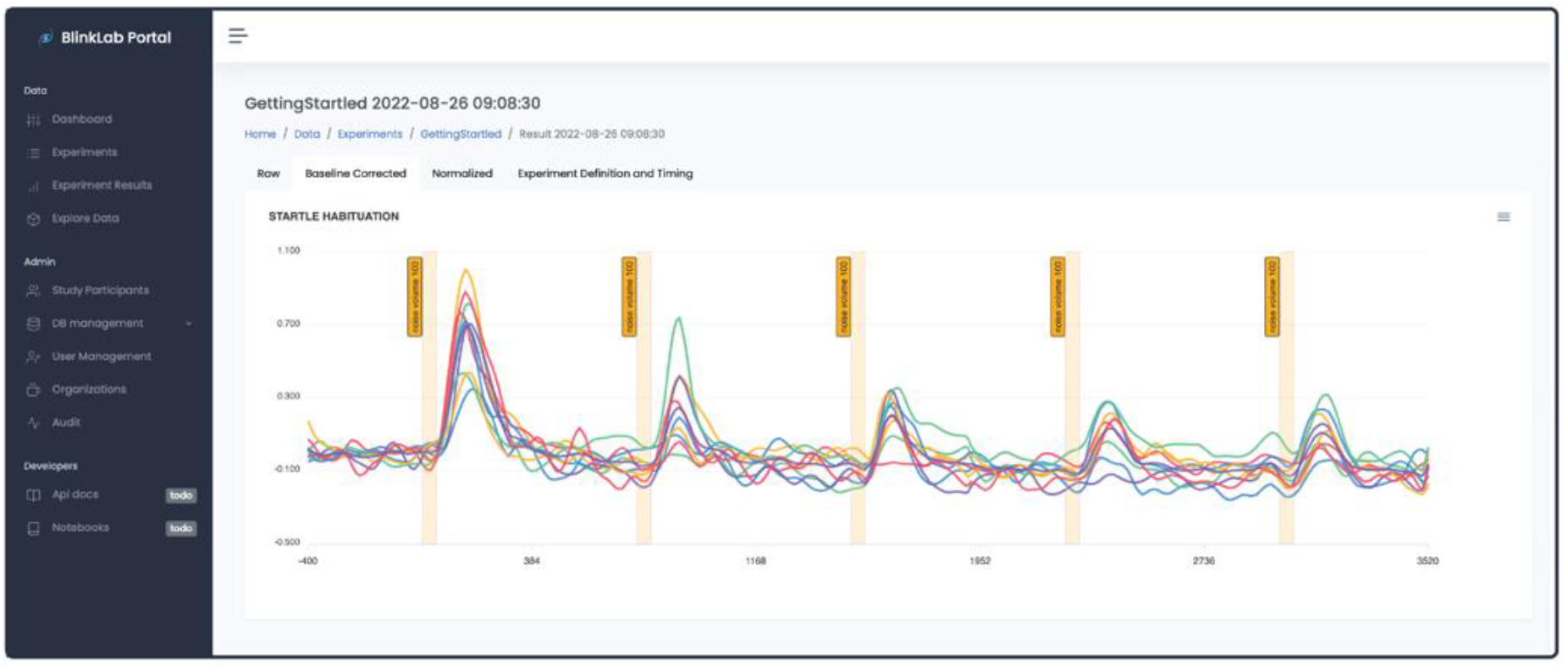
Raw eyelid traces of one participant for one startle habituation experiment. This figure is a screen capture of the cloud-based analysis portal and shows how we visualize the data that is collected with the smartphone. Each colored line represents one trial. Five consecutive startle pulses at 105 dB were presented with an interval of 750 ms between the trials. Note the gradual decrease in eyelid startle responses.

**Supplementary table 1.** Participants per neurometric test.

|  | **Eyeblink**<br>**conditioning**<br>**(n = 14)** | **Prepulse**<br>**inhibition**<br>**(n = 30)** | **Startle habituation**<br>**(n = 14)** |
| --- | --- | --- | --- |
| **Age (years)\*** | 29 (± 13.17) | 25 (± 13.82) | 34 (± 17.87) |
| **Sex** |  |  |  |
| Male | 5 (36%) | 14 (47%) | 6 (43%) |
| Female | 9 (64%) | 16 (53%) | 8 (57%) |

*\* All values: mean ± 1 standard deviation*

**Supplementary table 2.** Pavlovian eyeblink conditioning (n = 14 participants)

| Session | Phase | ISI duration | CS duration |  | CR percentage<br>(mean ± 95% CI) | CR amplitude<br>(mean ± 95% CI) | Latency to CR onset (ms)<br>(mean ± 95% CI) | Latency to CR peak (ms)<br>(mean ± 95% CI) |
| --- | --- | --- | --- | --- | --- | --- | --- | --- |
|  |  |  | CS-US | CS only |  |  |  |  |
| 0 | Baseline | 400 ms | 450 ms | 450 ms | 5.3 (± 6.6) | -0.02(± 0.05) | 156.33(± 121.24) | 229(± 115.18) |
| 1 | Acquisition | 400 ms | 450 ms | 450 ms | 20.4 (± 10.0) | 0.08(± 0.07) | 163.47(± 34.78) | 412.5(± 96.4) |
| 2 | Acquisition | 400 ms | 450 ms | 450 ms | 31.0 (± 12.5) | 0.15(± 0.08) | 179.11(± 27.3) | 531.96(± 31.71) |
| 3 | Acquisition | 400 ms | 450 ms | 450 ms | 37.0 (± 13.1) | 0.21(± 0.1) | 167.19(± 29.9) | 436.69(± 60.53) |
| 4 | Acquisition | 400 ms | 450 ms | 450 ms | 49.3 (± 13.0) | 0.3(± 0.11) | 183.24(± 27.97) | 455.63(± 31.51) |
| 5 | Acquisition | 400 ms | 450 ms | 450 ms | 56.6 (±11.1) | 0.36(± 0.1) | 170.41(± 28.7) | 492.65(± 25.28) |
| 6 | Acquisition | 400 ms | 450 ms | 450 ms | 58.5 (10.6) | 0.35(± 0.09) | 203.55(± 26.47) | 470.46(± 26.53) |
| **Main effect session\*** |  |  |  |  | F(6,4044) = 65.13, p < .00001 | F(6,4044) = 74.82, p < .00001 | F (6,1057) = 1.65, p = 0.13 | F (6,270) = 8.92, p = 6.79E-09 |
| 6 | Short CS | 400 ms | 450 ms | 450 ms | 58.46 (± 10.55) | 0.35 (± 0.09) | 203.55 (± 26.47) | 470.46 (± 26.53) |
| 7 | Short CS | 400 ms | 450 ms | 100 ms | 55.77 (± 14.55) | 0.34 (± 0.11) | 172.81 (± 28.47) | 475.53 (± 37.64) |
| **Main effect session\*** |  |  |  |  | F(1,1169) = 0.57, p=0.45 | F(1,1169) = 0.68, p=0.41 | F(1,387) = 3.64, p=0.06 | F(1,82) = 0.42, p=0.52 |
| 6 | ISI switch | 400 ms | 450 ms | 450 ms | 58.46 (± 10.55) | 0.35 (± 0.09) | 203.55 (± 26.47) | 470.46 (± 26.53) |
| 8 | ISI switch | 700 ms | 750 ms | 750 ms | 30.58 (± 15.43) | 0.14 (± 0.15) | 236.78 (± 31.24) | 652.97 (± 91.6) |
| 9 | ISI switch | 700 ms | 750 ms | 750 ms | 40.23 (± 18.71) | 0.22 (± 0.18) | 278.32 (± 78.02) | 765.6 (± 75.85) |
| **Main effect session\*** |  |  |  |  | F(2,1522) = 65.8, p < .00001 | F(2,1522) = 56.22, p < .00001 | F(2,749) = 3.42, p=0.03 | F(2,113) = 42.21, p < .00001 |

*All values: mean ± 95% confidence interval*
*\* ANOVA on Linear Mixed-Effect model*

**Supplementary table 3.** Prepulse inhibition of acoustic startle response (n = 30 participants)

| **Trial type** | **Amplitude of eyelid startle in response to pulse** |  |  | **Amplitude of eyelid startle in response to prepulse** |  |  |
| --- | --- | --- | --- | --- | --- | --- |
| Pulse (105 dB) | 0.35 (± 0.08) | | | 0.01 (± 0.02) | | |
| Prepulse 65 dB + Pulse | 0.14 (± 0.06) | | | 0.01 (± 0.03) | | |
| Prepulse 75 dB + Pulse | 0.1 (± 0.05) | | | 0.02 (± 0.02) | | |
| Prepulse 83 dB + Pulse | 0.09 (± 0.04) | | | 0.05 (± 0.03) | | |
| Prepulse 93 dB + Pulse | 0.07 (± 0.03) | | | 0.1 (± 0.03) | | |
| Main effect of trial type* | F(4,1434) = 99.44, p<.00001 |  |  | F(4,1434) = 15.62, p<.00001 |  |  |

| **Pairwise differences of trial type** | **Estimate** | **t-ratio** | **p-value\*\*** | **Estimate** | **t-ratio** | **p-value\*\*** |
| --- | --- | --- | --- | --- | --- | --- |
| (Pulse) vs. (PP 65 dB + Pulse) | 0.203 | 12.758 | <.0001 | -0.0097 | -0.724 | 1 |
| (Pulse) vs. (PP 75 dB + Pulse) | 0.249 | 15.505 | <.0001 | -0.0104 | -0.771 | 1 |
| (Pulse) vs. (PP 83 dB + Pulse) | 0.258 | 15.873 | <.0001 | -0.0477 | -3.483 | 0.0036 |
| (Pulse) vs. (PP 93 dB + Pulse) | 0.279 | 17.291 | <.0001 | -0.0912 | -6.701 | <.0001 |
| (PP 65 dB + Pulse) vs. (PP 75 dB + Pulse) | 0.046 | 2.913 | 0.0145 | -0.0007 | -0.055 | 1 |
| (PP 65 dB + Pulse) vs. (PP 83 dB + Pulse) | 0.054 | 3.423 | 0.0032 | -0.0380 | -2.82 | 0.0244 |
| (PP 65 dB + Pulse) vs. (PP 93 dB + Pulse) | 0.075 | 4.771 | <.0001 | -0.0814 | -6.085 | <.0001 |
| (PP 75 dB + Pulse) vs. (PP 83 dB + Pulse) | 0.008 | 0.535 | 0.5926 | -0.0372 | -2.739 | 0.025 |
| (PP 75 dB + Pulse) vs. (PP 93 dB + Pulse) | 0.029 | 1.851 | 0.1931 | -0.0807 | -5.973 | <.0001 |
| (PP 83 dB + Pulse) vs. (PP 93 dB + Pulse) | 0.021 | 1.298 | 0.3889 | -0.0434 | -3.181 | 0.009 |

*All values: mean ± 95% confidence interval*
*\* ANOVA on Linear Mixed-Effect model*
*\*\*P-value adjustment: bonferroni-holm method for 10 tests*

**Supplementary Table 4.** Startle habituation (n = 14 participants)

| Pulse number | Percentage eyelid startles |  |  | Eyelid startle amplitude |  |  |
| --- | --- | --- | --- | --- | --- | --- |
| Startle pulse 1 | 39.68 (± 22.47) | | | 0.30 (± 0.15) | | |
| Startle pulse 2 | 27.78 (± 20.4) | | | 0.16 (± 0.14) | | |
| Startle pulse 3 | 21.43 (± 20.05) | | | 0.13 (± 0.15) | | |
| Startle pulse 4 | 21.43 (± 21.12) | | | 0.11 (± 0.14) | | |
| Startle pulse 5 | 19.84 (± 19.19) | | | 0.13 (± 0.14) | | |
| **Main effect of pulse\*** | F(4,612) = 9.85, p<.00001 |  |  | F(4,612) = 18.14, p<.00001 |  |  |

| Pairwise differences | Estimate | t-ratio | p-value** | Estimate | t-ratio | p-value** |
| --- | --- | --- | --- | --- | --- | --- |
| Startle pulses 1 vs. 2 | 11.9 | 3.215 | 0.0096 | 0.135 | 5.459 | <.0001 |
| Startle pulses 1 vs. 3 | 18.25 | 4.93 | <.0001 | 0.165 | 6.7 | <.0001 |
| Startle pulses 1 vs. 4 | 18.25 | 4.93 | <.0001 | 0.182 | 7.387 | <.0001 |
| Startle pulses 1 vs. 5 | 19.84 | 5.358 | <.0001 | 0.164 | 6.665 | <.0001 |
| Startle pulses 2 vs. 3 | 6.35 | 1.715 | 0.4345 | 0.030 | 1.242 | 1 |
| Startle pulses 2 vs. 4 | 6.35 | 1.715 | 0.4345 | 0.047 | 1.928 | 0.325 |
| Startle pulses 2 vs. 5 | 7.94 | 2.143 | 0.1949 | 0.029 | 1.207 | 1 |
| Startle pulses 3 vs. 4 | 0 | 0 | 1 | 0.017 | 0.687 | 1 |
| Startle pulses 3 vs. 5 | 1.59 | 0.429 | 1 | -0.000 | -0.035 | 1 |
| Startle pulses 4 vs. 5 | 1.59 | 0.429 | 1 | -0.017 | -0.722 | 1 |

*All values: mean ± 95% confidence interval*
*\* ANOVA on Linear Mixed-Effect model*
*\*\*P-value adjustment: bonferroni-holm method for 10 tests*

## Notes

### Competing Interest Statement

Henk-Jan Boele, Sebastiaan Koekkoek, Peter Boele, Samuel S.-H. Wang, Anton Uvarov, and Chris de Zeeuw engage with BlinkLab Pty Ltd, the exclusive licensee of the technology, as co-founders and equity holders. The remaining authors declare no competing interests.

### Funding Statement

Henk-Jan Boele and Sam Wang were sponsored by the Princeton Accelerator Grant 2021

### Author Declarations

All procedures were approved by both the Institutional Review Board for Human Subjects of Princeton University (IRB#13943) and the Medical Ethics Review Committee of Erasmus MC (# MEC- 2022-36 0116).

